## Supplementary tabel 1 for "The impact of colectomy and chemotherapy on risk of type 2 diabetes onset in patients with colorectal cancer: Nationwide cohort study in Denmark"

**Supplementary tabel 1**: Likelihood of developing type 2 diabetes after different types of colorectal cancer surgery with and without oncological treatment, adjusted analysis

| **Supplementary table 1. Likelihood of developing type 2 diabetes after different types of colorectal cancer surgery with and without oncological treatment, adjusted for UICC alone and in Model 1 and Model 2** | | | | | | |
| --- | --- | --- | --- | --- | --- | --- |
|  | Adjusted for UICC | | Model 1 + UICC | | Model 2+UICC | |
|  | HR (95%CI) | *p-value* | HR (95%CI) | *p-value* | HR (95%CI) | *p-value* |
| **Surgery and chemotherapy** |  |  |  |  |  |  |
| Right-No-Chemo | ref |  |  |  |  |  |
| Right-Chemo | 0.98 (0.83;1.15) | 0.788 | 0.93 (0.78;1.11) | 0.441 | 0.97 (0.81;1.16) | 0.731 |
| Left-No-Chemo | 1.11 (0.98;1.24) | 0.097 | 0.93 (0.81;1.06) | 0.269 | 0.94 (0.83;1.07) | 0.374 |
| Left-Chemo | 1.03 (0.89;1.19) | 0.730 | 0.88 (0.74;1.03) | 0.120 | 0.93 (0.79;1.09) | 0.373 |
| Rectal-No-Chemo | 0.93 (0.82;1.05) | 0.225 | 0.81 (0.71;0.93) | 0.002 | 0.85 (0.74;0.97) | 0.018 |
| Rectal-Chemo | 0.89 (0.76;1.04) | 0.139 | 0.76 (0.64;0.90) | 0.002 | 0.82 (0.69;0.97) | 0.022 |
| Right: Right-sided colonic resections; Left: Left-sided colonic resections; Rectal: Rectal resections  No-Chemo: No chemotherapy; Chemo: Chemotherapy  All Colon: All right- and left-sided colonic resections with and without chemotherapy  All Rectal: All rectal resections with and without chemotherapy, and with and without radiation therapy  Model 1: adjusted for year of surgery, age at surgery, sex, BMI  Model 2: adjusted like Model 1 and further adjusted for performance status before surgery assessed by American Society of Anesthesiologist physical scale, smoking and alcohol | | | | | | |
